# Structural variation in nebulin and its implications on phenotype and inheritance: establishing a dominant distal phenotype caused by large deletions

**DOI:** 10.1101/2024.10.04.24313542

**Authors:** Lydia Sagath, Kirsi Kiiski, Kireshnee Naidu, Krutik Patel, Per Harald Jonson, Milla Laarne, Djurdja Djordjevic, Grace Yoon, Anna LaGroon, Curtis Rogers, Maureen Kelly Galindo, Katalin Scherer, Erdmute Kunstmann, Erkan Koparir, Desirée Ho, Mark Davis, Purwa Joshi, Alexander Zygmunt, Rotem Orbach, Sandra Donkervoort, Carsten G. Bönnemann, Marco Savarese, Andoni Echaniz-Laguna, Valérie Biancalana, Casie A. Genetti, Susan T. Iannaccone, Alan H. Beggs, Carina Wallgren-Pettersson, Franclo Henning, Katarina Pelin, Vilma-Lotta Lehtokari

## Abstract

**Introduction:** Structural variants (SVs) of the nebulin gene (*NEB*), including intragenic duplications, deletions, and copy number variation of the triplicate region, are an established cause of recessively inherited nemaline myopathies and related neuromuscular disorders. Large deletions have been shown to cause dominantly inherited distal myopathies. Here we provide an overview of 35 families with muscle disorders caused by such SVs in *NEB*.

**Methods:** Using custom Comparative Genomic Hybridization arrays, exome sequencing, short-read genome sequencing, custom Droplet Digital PCR, or Sanger sequencing, we identified pathogenic SVs in 35 families with *NEB*-related myopathies.

**Results:** In 23 families, recessive intragenic deletions and duplications or pathogenic gains of the triplicate region segregating with the disease in compound heterozygous form, together with a small variant in trans, were identified. In two families the SV was, however, homozygous. Eight families have not been described previously. In 12 families with a distal myopathy phenotype, eight unique, large deletions encompassing 52 to 97 exons in either heterozygous (n = 10) or mosaic (n = 2) state were identified.

In the families where inheritance was recessive, no correlation could be made between the types of variants and the severity of the disease. In contrast, all patients with large dominant deletions in *NEB* had milder, predominantly distal muscle weakness.

**Discussion:** For the first time, we establish a clear and statistically significant association between large *NEB* deletions and a form of distal myopathy. In addition, we provide the hitherto largest overview of the spectrum of SVs in *NEB*.

## 1. Introduction

### 1.1. The nebulin gene

Nebulin (*NEB*, HGNC:7720, MIM ID *161650, genomic location 2q23.3) is a large protein expressed in the skeletal muscle sarcomere, integral to its structure and function. Its 183 exons span a genomic region over 250 kb, giving it a theoretical full-length mRNA of 26 kb, encoding more than 8,600 amino acids with a combined molecular weight of approximately 990 kDa.

Exons 63-66, 143-144, and 167-177 of *NEB* undergo alternative splicing, giving rise to various nebulin isoforms. ^1,2^ In addition, exons 82-105 constitute an intragenic segmental duplication region, referred to as the triplicate region (*NEB* TRI). This region consists of a 10 kb, eight-exon block repeated three times (exons 82-89, 90-97, 98-105), at 99% identity between the repeats; thus, the normal number of eight-exon blocks in a diploid genome is six. ^1^

The C-terminal end of nebulin encodes a Z-disc anchor, while the N-terminus is located in the pointed cap end of the thin filament. ^3,4^ The full-length protein consists of 29 actin-binding super repeats, each composed of seven simple actin-binding motifs (SDxxYK), allowing the protein to potentially bind to more than 200 actin monomers. The super repeats closer to the Z-disc and the cap end bind actin with a stronger affinity than the super repeats in the middle of the protein. ^5^ Each *NEB* TRI eight-exon block encodes for two super repeats, resulting in six super repeats in total in the wildtype allele TRI region. ^6^ It was previously hypothesized that each super repeat also contains a tropomyosin-binding site (WLKGIGW). ^1,3^ however, it has recently been shown that nebulin and tropomyosin do not directly bind to each other. ^7^ Instead, the WLKGIGW motif and an ExxK motif in nebulin seem to interact with the linker region of troponin T. ^7^ The large number of nebulin isoforms is thought to contribute to the regulation of thin filament length and Z-disc structure in different fiber types and under differing physiological conditions.^8-10^

Nebulin stiffens the thin filament and is critical for the generation of physiological force levels. ^11^ The thin filament can be assembled in the absence of nebulin, ^12,13^ but its absence results in shorter actin filaments, impaired tropomyosin/troponin movement on the thin filament, altered cross-bridge formation, and lesser force. ^11,14^

### 1.2. Variation in nebulin

To date, more than 800 variants in *NEB* have been recorded in the Leiden Open Variation Database v.3.0 (LOVD, April 25 2024), ^15^ and of these, 385 (46%) have been classified as pathogenic or likely pathogenic. This high number of variants can be explained by the size of the gene and the spontaneous mutation rate of the human genome. The majority (95%) of them are small variants; truncating variants (56%), variants affecting splicing (27%), missense variants (9%), and other types of small variants (2.5%).

The Genome Aggregation Database (gnomAD, August 14 2024) SVs 4.1.0 holds 30 records of SVs in *NEB* over 50 bp in size encompassing at least one exon. Eight of these are different variants of the *NEB* TRI region. The allele frequency of all of the recorded SVs is less than 0.01. The full data extracted from gnomAD SVs 4.1.0 is available in Supplementary Table 2, and more details about the data can be found in Supplementary File 1.

SVs in *NEB* may be divided into two categories: 1. *NEB* TRI copy number variants (*NEB* TRI CNVs), which refer to gains and losses limited to the *NEB* TRI region (1.5%), 2. deletions and duplications over 50 bp outside the *NEB* TRI region, referred to here as structural variants (SVs) (5%). These SVs form 5% of pathogenic variants in *NEB* in LOVD (Figure 1, Supplementary Table 1).

**Figure 1.**
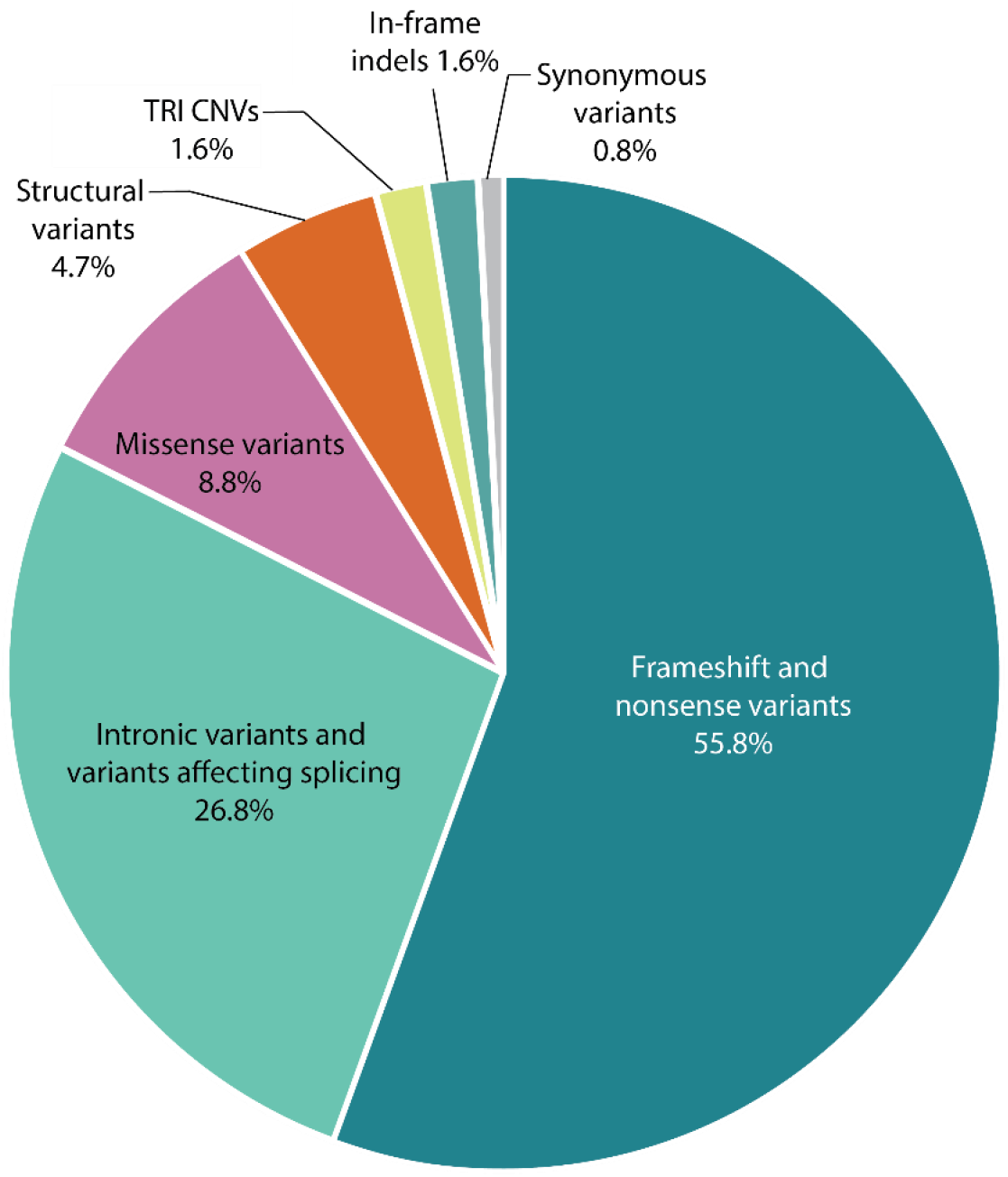
Distribution of pathogenic variants in NEB recorded in LOVD. Full data are available in Supplementary Table 1. Abbreviations: TRI = NEB triplicate region, exons 82-89, 90-97, 98-105; CNVs = copy number variants.

While the loss or gain of one of the three repeated *NEB* TRI blocks per allele is considered a benign copy number polymorphism, gains of two or more blocks per allele are recessive pathogenic CNVs. ^6^ No losses of more than one block on one allele have been recorded. CNVs of the *NEB* TRI region are seen in up to 14% of congenital myopathy samples, ^16,17^ and approximately half are pathogenic. ^18^ To our knowledge, no inversions or translocations in *NEB* have been recorded to date.

To date, all single nucleotide variants (SNVs), small indels, and TRI CNVs described are recessive, while SVs in *NEB* can be divided into recessive and dominant SVs. Recessive SVs are either frameshifting or null-alleles, while the previously described dominant deletions are in-frame, allowing the expression of stable mini-nebulin proteins. ^6,19-22^ In addition, a well-characterized recessive deletion of exon 55 is a known nemaline myopathy (NM)-causing founder mutation identified in people of Ashkenazi Jewish population (prevalence 1:108). ^23^

### 1.3. The nebulin gene and disease – a spectrum of myopathies

Pathogenic variants in *NEB* were first described in patients with congenital autosomal recessive (AR) NM type 2 (NEM2, MIM ID #256030), ^8^ but have since been found to cause other related congenital myopathies as well. Pathogenic variants in *NEB* are the most common cause of recessive NM^8,23-30^ and are responsible for roughly half of all cases. ^31,32^ NM, in turn, is one of the most common of the rare congenital myopathies with a birth incidence of ∼1 in 50,000 live births. ^31^

NMs are highly variable in severity, usually presenting as skeletal muscle weakness of early onset, predominantly present in the proximal limb, respiratory, facial, and trunk muscles, with a later distal involvement. The patients run a risk of insidious decrease in respiratory function. A hallmark identifier is ‘nemaline bodies’ in Gömöri trichrome-stained muscle biopsy sections. ^33^ These protein aggregates consist of accumulations of Z-disc and thin filament proteins. NEM2 can be caused by compound heterozygous or homozygous SNVs and indel variants in *NEB*, partial deletions or duplications of *NEB*, and copy number variants (CNVs) of the *NEB* TRI region. In addition, recessive *NEB* variants have been identified in patients with congenital myopathy with cap-like structures and nemaline bodies, ^34^ core-rod myopathy, ^35^ and lethal neonatal arthrogryposis multiplex congenita-6 (AMC6, MIM ID #619334). ^36^ Distal forms of recessive myopathies with or without nemaline bodies, and core-rod myopathy have been reported, varying in age of onset, and caused by biallelic *NEB* variants, have been reported.^19,20,37-40^

The same recessive *NEB* variant may in one patient be associated with NM, but in another patient, in combination with another recessive variant, cause another, related congenital myopathy. ^19,20,34,36^ Variants in constitutional exons are most often associated with typical forms of NM, while variants in alternatively spliced exons are more common in patients with other or more unusual forms of NM. ^19^

Two families with distal phenotypes as a result of large dominant deletions in *NEB* have previously been described. ^21,22^ One of these families had a dominantly inherited large deletion segregating in three generations, and in the second family, the index patient had a large *de novo* deletion in mosaic form. These patients with distal forms of NEB-related myopathies did not have decreased respiratory function, and in general the phenotype was milder.

In this article, we describe ten new families with distal phenotypes caused by six different large in-frame deletions, significantly expanding the number of large structural dominant variants identified in *NEB*. One of the new deletions presented existed in mosaic form. All these large dominant deletions were associated with a distal phenotype. We also present two new families with congenital myopathies caused by recessive SVs in *NEB* in combination with small variants, and five new families with pathogenic CNVs of the *NEB* TRI region. In addition, we review the published cases with large *NEB* SVs and CNVs of the *NEB* TRI region.

## 2. Materials & Methods

### 2.1. Samples

Altogether DNA samples from 35 families, and muscle biopsies from 3 families, were included in this study. The patient samples used were heterogeneous in terms of clinical diagnosis, ethnic background, and age. Altogether 22 families had a NM diagnosis: three of these classified as severe, seven as typical, one as mild, three as other/unusual forms, and in nine families the classification was unknown. In addition, the index patient of one family had core-rod myopathy. Milder distal myopathy was the primary diagnosis in 12 families.

The DNA stocks were extracted either from peripheral blood, saliva, or muscle biopsy. DNA was eluted into TE buffer or water, and stored at -20°C or -80°C. The DNA concentration and quality were measured using DeNovix DS-11 FX+ Spectrophotometer/Fluorometer (DeNovix Inc., Wilmington, DE, USA). Subsequent dilutions were done in sterile water in appropriate series.

### 2.2. Genetic analysis methods

SVs and *NEB* TRI CNVs were identified and validated using different methods as described below. The samplewise methods are listed for samples with *NEB* TRI CNVs in Supplementary Table 4, for recessive *NEB* SVs in Supplementary Table 5, and for dominant *NEB* SVs in Supplementary Table 6. All *NEB* cDNA variants are identified relative to reference transcript NM_001271208.2, corresponding to protein reference sequence NP_001258137.2.

#### 2.2.1. Comparative Genomic Hybridization array

Altogether DNA samples of 96 persons (50 healthy and 46 affected) from 33 families were run on a custom 4×180k or 8×60k comparative genomic hybridization array (NM-or NMD-CGH-array) as previously described. ^16,17^ The degree of mosaicism of the patients with deletions in mosaic state (F24, F33) was determined as previously described. ^22^

#### 2.2.2. Droplet digital PCR

For eleven samples from eight families with *NEB* TRI CNVs (F1, F3-F4, F7-F8, and F10-F12), custom droplet digital PCR (ddPCR) assays for the *NEB* TRI region were run as previously described. ^18^

#### 2.2.3. Sequencing

The sequencing methods used included different short-read gene panels, exome sequencing (ES), short-read genome sequencing (GS), and Sanger sequencing.

Samples from families F26-F30 were subjected to a search for SNV/small indel identification protocol as previously described. ^41^ Subsequently, uncharacterized patient samples underwent CNV detection via a multi-method pipeline, which used cn.mops, ^42^ clinCNV, ^43^ and ExomeDepth^44^ to explore potential CNVs. Filtration using the variant effect predictor (VEP) score^45^ and control population frequencies (gnomAD and DGVgold) were used to remove known benign variants. Finally, the SARC R package^46^ was used to remove detected CNVs with a low confidence score. Other SV types than CNVs were not possible to analyze from this data.

For F35, CANOES^47^ was used to generate CNV calls from ES data.

Of the previously unpublished families, F23, F26-F27, and F31-F34 had undergone commercial or other diagnostic short-read sequencing for which details are not available.

### 2.4. Protein analysis

In two families (F24, F25), muscle-derived proteins were analyzed on western blot to study nebulin isoform expression, as previously described. ^21^ In F35, muscle-derived proteins were analyzed on western blot as described by Warren and colleagues^48^, using mouse moloclonal antibodies against the nebulin super repeat 21, clone 9C4 (provided by Prof. Glenn Morris, Oswestry, UK) at 1:300 dilution and detected by enhanced chemiluminescence (ECL) (Supplementary Figure 1).

### 2.5. Statistical methods

All statistical tests were performed in RStudio 2022.02.1 build 461. Details concerning all statistical analyses are available in Supplementary File 1.

#### 2.5.1. Correlation analysis between gene length and number of recorded SVs

The genomic gene length and number of structural variants recorded in gnomAD SVs 4.1.0 were extracted for 13 known NM-causing genes (Supplementary Table 3). The gene length was plotted against the number of recorded SVs using ggplot2 to verify normal distribution. The Pearson correlation coefficient and p-value were calculated.

#### 2.5.2. Correlation analysis between different types of small variants and phenotype

Families with recessive variants and sufficient phenotypic data (n = 16) were classified into three groups: mild (n = 3), typical (n = 10), and severe (n = 3), according to established classification criteria. ^49^ Small variants were divided into splicing (n = 2), missense (n = 1), in-frame deletions (n = 1), and truncating variants (n = 12), of which the latter includes both nonsense and frameshift variants. Families with homozygous *NEB* SVs (F20) and TRI CNVs (F12), and families in which severity classification was not possible (F2, F5, F13, F17, F19) were excluded from the statistical test. The correlation between the types of variants and the severity of the phenotype was tested with Fisher’s exact test.

#### 2.5.3. Correlation analysis between pathogenic NEB TRI CNV allele and phenotype

In families with pathogenic gains of the *NEB* TRI region, the correlation between the copy number of the *NEB* TRI of the pathogenic allele and the severity of phenotype was tested using Kendall’s Tau.

#### 2.5.4. Difference in deletion length between families with NM and distal phenotypes

The length of all unique deletions, both in recessive NM families (n = 7) and dominant distal myopathy families (n = 8), was extracted both in terms of number of affected exons and number of base pairs deleted. For deletions in which the exact breakpoints were unknown, the average length based on the CGH-array variant call was calculated by using the longest and shortest lengths indicated by the variant call in HGVS format. Data normality was assessed using the Shapiro-Wilk test, the homogeneity of variances was assessed *post hoc* using Levene’s test, and the Wilcoxon rank-sum test was used to compare the number of exons deleted and the length in basepairs between the recessive and dominantly segregating deletions. The difference between the groups was also assessed by an independent samples t-test.

## 3. Results

### 3.1. *NEB* TRI CNVs

We present 14 patients from 12 families (F1-F12) in whom pathogenic CNVs in the *NEB* TRI region were detected, five of which are presented here for the first time. In one family (F12), the index patient was homozygous for a pathogenic TRI CNV (7+7). The other index cases were compound heterozygous for a *NEB* TRI CNV in combination with another type of pathogenic variant (Table 1). The *NEB* TRI CNVs were identified using the NM-or NMD-CGH array in all 12 families, and the result was confirmed using custom ddPCR in eight families. The primary diagnosis of the patients with NEB TRI CNVs was NM confirmed by histopathological analyses and the severity of the disorder varied from mild to severe (Supplementary Table 4).

**Table 1.** Families with pathogenic NEB TRI CNVs. The complete variant details and additional information are available in Supplementary Table 4.

| Fam | Class | NEB TRI CNV |  |  |  | 2nd pathogenic *NEB* variant |  |  |  |  |
| --- | --- | --- | --- | --- | --- | --- | --- | --- | --- | --- |
|  |  | CN | Mat. allele CN | Pat. allele CN | Pub. | Exon | cDNA | Protein change | Allele | Pub. |
| F1 | 3 | 11 | 7* | 4 | U | 3 | c.82del | p.(Ile28*) | Pat | U |
| F2 | 0 | 10 | 7* | 3 | <sup>6</sup> | 17i | c.1569+1G>A | p.(spl) | Pat | U |
| F3 | 3 | 10 | 7* | 3 | <sup>6</sup> | 28 | c.2784del | p.(Asp929Ilefs*28) | Pat | <sup>6, 25</sup> |
| F4 | 2 | 14 | 11* | 3 | U | 59i | c.8256+1G>A | p.(spl) | Pat | U |
| F5 | 0 | 10 | N/A | N/A | <sup>6</sup> | 61i | c.8685+1G>A | p.(spl) | N/A | <sup>6, 19</sup> |
| F6 | 2 | 9 | 6* | 3 | <sup>6</sup> | 78 | c.11771_11788del | p.(Val3924_Asn3929del) | Pat | <sup>6, 25</sup> |
| F7 | 2 | 10 | 3 | 7* | <sup>6</sup> | 81 | c.12048_12049del | p.(Lys4017Argfs*10) | Mat | <sup>6</sup> |
| F8 | 2 | 10 | 7* | 3 | U | 126 | c.19712_19716delinsGAG | p.(Lys6507Argfs*4) | Pat | U |
| F9 | 2 | 10 | N/A | N/A | U | 158 | c.22936C>T | p.(Arg7646*) | N/A | U |
| F10 | 1 | 9 | 3 | 6* | <sup>6</sup> | 134 | c.20446del | p.(Arg6816Glyfs*38) | Mat | <sup>6, 19</sup> |
| F11 | 2 | 9 | 3 | 6* | <sup>6</sup> | 180 | c.25206T>A | p.(Tyr8402*) | Mat | U |
| F12 | 2 | 14 | 7* | 7* | U |  |  |  |  | U |

Fam: Family, Class 0: Unclassified/not known, 1: Mild, 2: Typical, 3: Severe, CN: Copy number, \*: Pathogenic allele, Mat: Maternal, Pat: Paternal, i: intron, Pub.: Published in, U: Previously unpublished.

### 3.2. Recessive SVs

Ten unique recessive SVs in *NEB* have been identified in 11 families (F13-F23). In nine families the patient’s SV was *in trans* with pathogenic SNVs or indels, while in F20 two affected siblings had the SV in homozygous form. The deletions and the second causative variants (SNVs or small indels) of the families and their respective associated phenotypes are presented in Table 2 and the SVs are visualized in Figure 2. An extended overview including detection and verification methods and more variant and phenotype details is provided in Supplementary Table 5. As the families with the recessive exon 55 deletion^50^ have previously been thoroughly described we do not present them in this article.

**Table 2.**
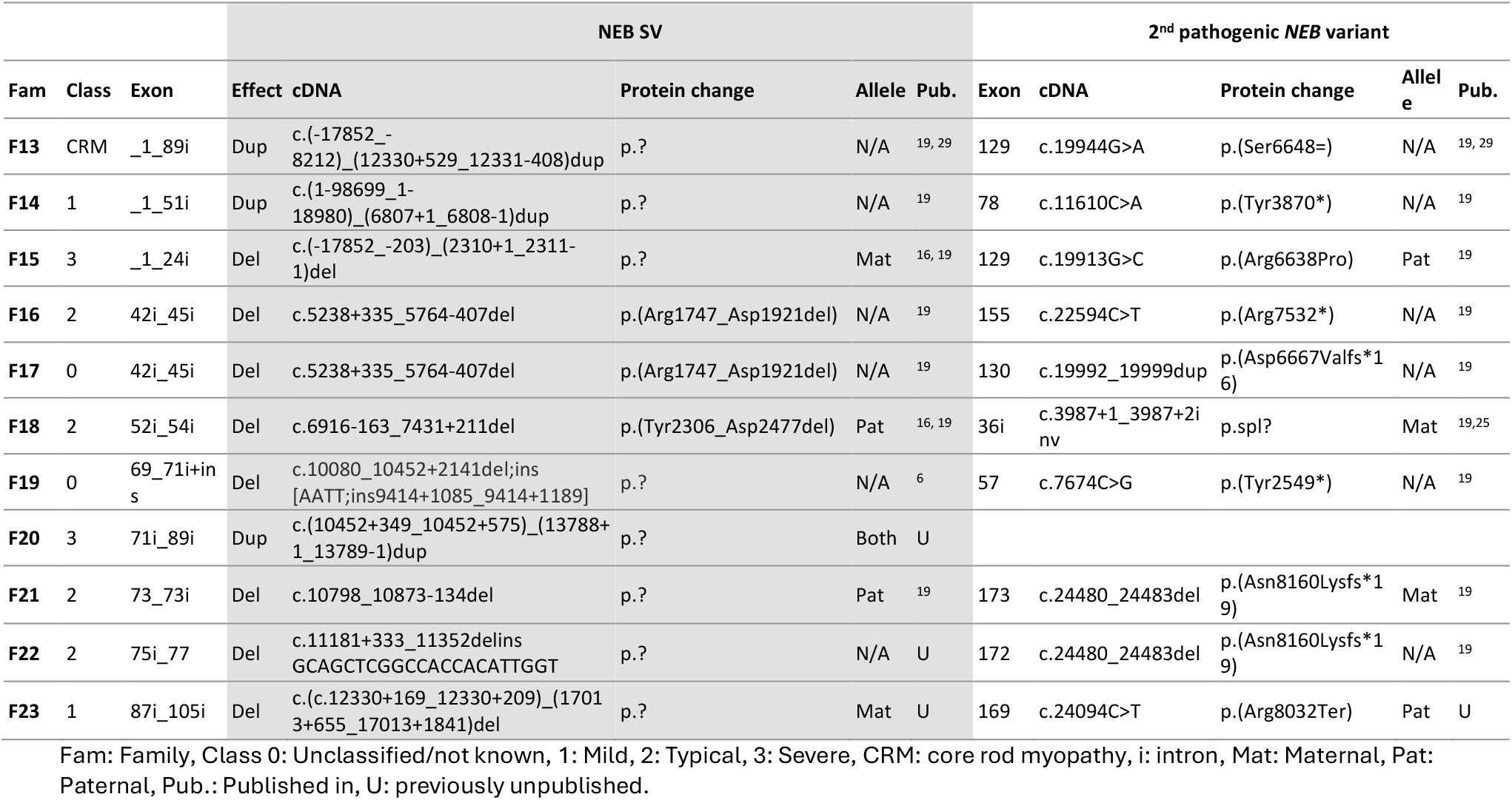
Families in which the causative variants are a combination of a recessive NEB SV and a pathogenic SNV or indel. The complete variant details are available in Supplementary Table 5.

**Figure 2.**
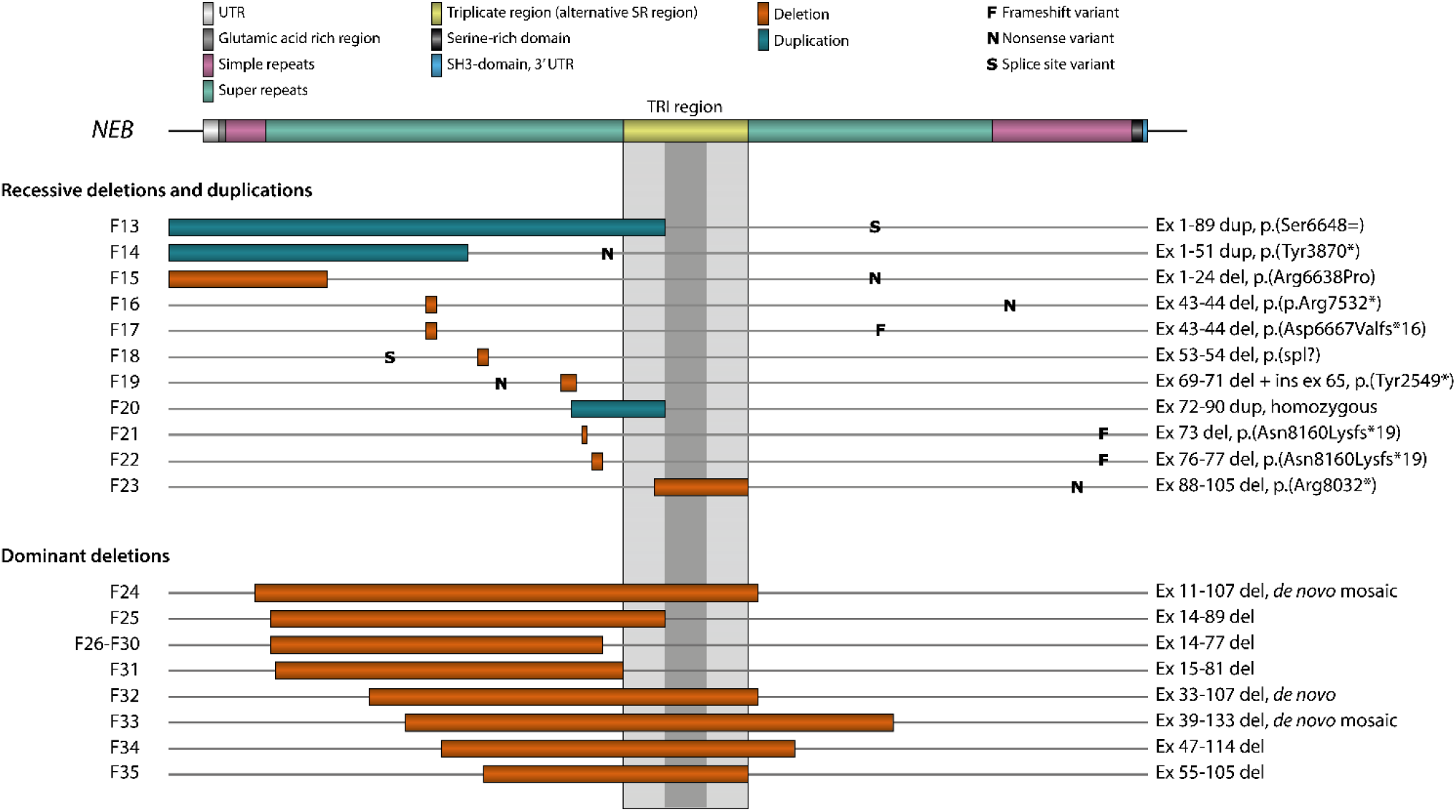
The structure of NEB, recessive SVs, and dominant deletions. CNVs of the NEB TRI region are not included in the figure. Detailed descriptions of the affected exons are listed in Tables 1-3.

The patients in all the families presented here, except one with recessive *NEB* SVs, had been diagnosed with NM, ranging from mild to severe forms of the disorder. In two families the phenotype could not be classified (F17 and F19), and in one family the muscle biopsy findings were consistent with core-rod myopathy (F13). The primary diagnoses of the patients with NM were confirmed by histopathological analyses and the severity of the disorder varied from mild to severe.

The largest recessive SV in *NEB* characterized to date is a deletion of exons 1-24 in F15, a patient with the severe form of NM. In addition, three different in-frame deletions (deletions of exons 43-45, 53-54, and 88-105) in four families (F16-F18 and F23) ranging from 2 to 18 exons in size have been identified. Two families (F16 and F17) share the same in-frame deletion of exons 43-45, but have different truncating *NEB* variants on the other allele. These families are seemingly unrelated but have the same country of origin.

Frameshift deletions affecting 1-3 exons leading to premature truncation of *NEB* were identified in families F19, F21, and F22. The second variants in these three families were also truncating.

In three families (F13, F14, F20), in-frame duplications of 18 to 89 exons in size were detected. The families with heterozygous duplications of exons 1-89 (F13) and 1-51 (F14) have been previously described ^19,29^. These patients had mild forms of NM (F14) and core-rod myopathy (F13). A previously unpublished duplication of exons 72-89 (F20) was found in homozygous form in two siblings in a consanguineous family, both affected by the severe form of NM.

In 9 out of 11 families, the SV was identified using the NMD-CGH-array, and in two families the preliminary finding was made by ES. For the latter, the breakpoints were subsequently verified using the NMD-CGH-array. In seven families the exact variant breakpoints were verified by Sanger sequencing as well, and in one, the finding was verified by multiplex ligation-dependent probe amplification (MLPA).

### 3.3. Dominant deletions

Large dominant *NEB* deletions have previously been reported in two separate families (F24 and F25) with distal nebulin myopathy. Here, we report 10 new families (F26-35) with distal myopathy caused by dominant deletions ranging from 52 to 98 exons in size (Table 3), all shown or predicted to be in-frame. The deletions are visualized in Figure 2.

**Table 3.** Dominant deletions in families with distal myopathies. The complete variant details are available in Supplementary Table 4.

| Fam | Affected/number of family members | Exon | cDNA | Protein | Inheritance | Pub. |
| --- | --- | --- | --- | --- | --- | --- |
| F24 | 1/3 | 10i_107i | c.(823_17014)=/del | p.(Gln275_Asn5671del) | De novo mosaic | 22 |
| F25 | 3/13 | 13i_89i | c.(1257+1_1258-1)_(13788+1_13789-1)del | p.(Lys420_Asp4596del) | AD familial | 21 |
| F26 | 11/21 | 13i_77i | c.(1152+1191_1153-550)_(11601+48_11601+862)del | p.? | AD familial | U |
| F27 | 11/29 | 13i_77i | c.(1152+1_1153-1)_(11601+1_11602-1)del | p.? | AD familial | U |
| F28 | 8/36 | 13i_77i | c.(1152+1191_1153-550)_(11601+48_11601+862)del | p.? | AD familial | U |
| F29 | 1/23 | 13i_77i | c.(1152+1191_1153-550)_(11601+48_11601+862)del | p.? | N/A | U |
| F30 | 12/29 | 13i_77i | c.(1152+1_1153-1)_(11601+1_11602-1)del | p.? | AD familial | U |
| F31 | 4/19 | 14i_81i | c.(1152+1_1153-1)_(12330+1_12331-1)del | p.? | AD familial | U |
| F32 | 1/1 | 32i_107i | c.(3256-53_3256-43)_(17013+655_17013+1841)del | p.? | De novo | U |
| F33 | 1/3 | 39_133i | c.(4507-9_4518)_(20367+433_20367+453)=/del | p.? | De novo mosaic | U |
| F34 | 4/5 | 46i_114i | c.(5971-491_5971-74)_(18156+42_18156+62)del | p.(His1991_Asp6052del) | AD familial | U |
| F35 | 4/10 | 54i_105i | c.(7431+1203_7431+1310)_(16705-481_16705-87)del | p.(Arg2478_Asp5568del) | AD familial | U |

Fam: family, i: intron, AD: autosomal dominant, Pub.: Published in, , i: Kiiski & Lehtokari et al. 2019, j: Sagath & Lehtokari et al. 2021, U: Previously unpublished.

In the previously reported family F25, a deletion of exons 14-89 was identified in three generations. In F24 the *de novo* deletion of exons 11-107 was identified in mosaic form in a patient with asymmetry in muscle strength. Using western blot of muscle biopsy material, the allele with the deletion was shown to be expressed and to result in a significantly shorter nebulin protein product^21,22^ in affected members of both of these families.

In two of the previously unreported families, the deletion identified had occurred *de novo*. In F32, the heterozygous deletion of exons 33-107 (75 exons) was initially identified by ES and subsequently confirmed using the NMD-CGH-array. In F33, a patient presenting with asymmetry in muscle strength, the deletion of exons 39-133 (95 exons) was initially identified through a gene panel, and the finding was confirmed using the NMD-CGH-array, showing a degree of mosaicism of 60% in DNA extracted from blood. RNA sequencing of skeletal muscle mRNA from this patient’s diagnostic biopsy confirmed splicing of exon 38 to 134 in roughly half the transcripts (data not shown).

In eight families, the pathogenic deletion is seen in more than one family member, segregating with the disease in a dominant fashion. In five families (F26, F27, F28, F29, F30), a deletion of exons 14 -77 (64 exons) was first identified either by a gene panel analysis or ES. In three of these, the deletion was also confirmed using the NMD-CGH-array. The families are not known to be related, but likely share a common ancestor; the families are subject for another publication in preparation. In family F31, a deletion of exons 14-81 (68 exons) was identified by ES in two patients, but not verified on the NMD-CGH-array, as no sample was available. It thus remains unclear whether the deletion extends further into the *NEB* TRI region. In family F34, a deletion of exons 47-114 (68 exons) was identified by short-read sequencing in the index patient, who presented with adult-onset muscle weakness. The deletion was subsequently shown to be present in their two affected children. In family F35, a deletion of exons 55-105 (51 exons) was confirmed in five patients presenting with mainly distal myopathy, with some proximal involvement and scapular winging.

All patients with large dominant deletions in *NEB* presented with mild phenotypes and primarily distal involvement, with facial and proximal involvement along with scapular winging present in some patients. All of these patients were ambulatory. While no nemaline bodies were reported, muscle fiber atrophy, fiber size abnormalities, type 1 fiber predominance, and/or central core regions have been noted in the histological analyses in families F24, F25, and F35.

### 3.4. Statistical analyses

The Pearson correlation coefficient between the gene length of NM-causing genes and number of recorded SVs in gnomAD SVs 4.1.0 was 0.982 (p = 2.534*10^−9^).

In this cohort, no statistically significant correlation was found between the type of small variant (truncating, splicing, or other) and the severity of the disorder in either the *NEB* TRI CNV families (p = 1) or *NEB* SV families (p > 0.05) separately, nor together (p > 0.05).

In contrast, a statistical significance was found between the size of the deletions segregating in a recessive versus a dominant fashion. The inheritance also correlated directly with the phenotype, where all dominant deletions were associated with a predominantly distal phenotype. Statistical significance was reached both in terms of the number of exons deleted (p = 0.0014) and the approximated size of the deletion (p = 0.0003), when grouped by phenotype or inheritance mode.

A correlation analysis between the length and number of exons affected in the groups of recessive SVs and dominant SVs suggests that dominant deletions are significantly larger than recessive deletions both in terms of the number of exons deleted and in terms of the length in base pairs (p < 0.0001 and p < 0.0001, Table 4).

**Table 4.** Number of exons and length in base pairs of recessive and dominant deletions in NEB, and statistical significance of differences between these groups.

| SV type | Recessive | Dominant | Difference significance |
| --- | --- | --- | --- |
| Number of unique deletions | 7 | 8 |  |
| Average number of exons (median; range) | 7.6 (2; 1-24) | 74.0 (72; 51-97) | $p = 1.961 \times 10^{-7}$ |
| Average length (median) | 13,461 bp (2506 bp) | 101,645.3 bp (97,640 bp) | $p = 2.21 \times 10^{-6}$ |

## Discussion

In this study, we describe 35 families presenting with either NM, NM-related disorders, or distal myopathies caused by SVs in *NEB* (Figure 2). These families constitute around 7% of the families included in our research database, now compromising over 520 families. The families included in this study can be divided into three groups: 12 families with recessive CNVs of the *NEB* TRI region, 11 families with recessive SVs (both duplications and deletions), and 12 families affected by large dominant deletions in heterozygous or mosaic form.

CNVs of the *NEB* TRI region constitute the most common recurrent form of benign SV in *NEB*. Benign or recessive CNVs of the *NEB* TRI region are present in all populations represented in gnomAD SVs v4.1.0, indicating that gains and losses of the *NEB* TRI show an independent recurrence. To date, the lowest *NEB* TRI copy number identified is 5 (3+2 per allele), and the highest 14 (in combinations of 11+3 and 7+7 per allele). The largest number of repeats on one allele recorded in our cohort is 11 (F3, copy number inferred by segregation). Benign losses of the *NEB* TRI region are marginally rarer than gains. ^18^

We have previously hypothesized that the pathogenetic mechanism behind gains of the *NEB* TRI region is based on the destabilization of mRNA or its secondary structure, thus affecting the translation process. ^6^ The alternative hypothesis has been that each gained block would add 486 amino acids and two super repeats to the protein, affecting the protein’s function as a stabilizer of the thin filament. In a recent study, Karimi & colleagues^51^ used RNA sequencing, agarose gel electrophoresis, and thin filament length measurements to show that only two gained blocks were expressed at the mRNA level in a patient with a four-copy gain of the *NEB* TRI region. Both hypotheses may thus hold; not all gained blocks seem to be transcribed, but a longer protein is expressed in addition to the wild-type ones. ^51^

Recessive SVs in *NEB*, both those previously published and those presented in this article, are either null alleles, truncating variants, or in-frame deletions of 1 to 24 exons (Anderson et al 2004, Lehtokari et al 2014). We hypothesize that all SVs encompassing the first three exons of the gene are recessive null variants, as the *NEB* start codon lies in exon 3; this applies to the largest recessive deletion in our cohort, a deletion of the 24 first exons of *NEB* (F15) and the large duplications of exons 1-89 (F13) and exons 1-51 (F14). No statistically significant genotype-phenotype correlations can be made among the disorders caused by recessive *NEB* variants.

Previously, all *NEB* variants, including SVs, were believed to be recessive. The first large, dominant deletions were described in 2019 and 2021. One of them segregated in a dominant fashion, and the other one was a *de novo* mosaic variant. ^21,22^ In this update, we present nine novel families with large heterozygous deletions and one (F33) with a large deletion in mosaic form. Both patients with mosaic deletions presented with distal asymmetric muscle weakness.

Large deletions seem to be consistently associated with predominantly distal phenotypes, clearly distinct from the NEM2 clinical presentation. Additionally, the patients with these large deletions are more likely to be mildly affected compared with the two other groups presented.

Expression analyses of these large in-frame deletions show expression of a shorter nebulin product and have previously been published for F24 and F25, ^21,22^ and was performed for F35 (Supplementary Figure 1). Histological findings in families with large dominant deletions are milder than in families with recessive NM and related disorders.

Currently known large recessive pathogenic deletions range from 1 to 24 exons, and duplications from 18 up to 89 exons in *NEB*. ^19,21,22^ The dominant deletions in our cohort vary in size between 51 to 97 exons, and all include the *NEB* exons 55 to 77. The upper limit of recessive deletion lengths is not currently known, but can possibly be deduced from the largest recessive and smallest dominant deletions in our cohort, and by the largest deletion recorded in gnomAD SVs 4.1.0, which spans ten exons (ex 159-168). We hypothesize that the threshold between a recessive and a dominant deletion may be anywhere between 24 and 51 exons.

Nebulin acts as a molecular ruler and stabilizer of the thin filament and contributes to physiological force generation. ^10,11^ As all large, disease-causing heterozygous deletions are in-frame, the most likely pathogenetic mechanism is that sufficiently large deletions disrupt sarcomere assembly and function through a dominant-negative effect due to the expression of a shorter protein product.

The N-terminal part of the protein constitutes the thin filament cap. The C-terminus differs from the other parts of the gene in coding for the Z-disc anchor. Additionally, more alternative splicing in this region has been observed in comparison with other parts of the gene. It has been hypothesized that the C-terminal part may be vital for the function of the protein and/or sarcomere structure. In our cohort, none of the SVs affect *NEB* exons 134-187, and all the deletions identified affect the super repeat-rich regions, including the *NEB* TRI region (in green and yellow in Figure 2). However, although our cohort lacks SVs located in the last third of the gene including the C-terminus, their presence in databases such as gnomAD SVs 4.1.0. indicates they are putative recessive pathogenic alleles. Whether a deletion is dominant or recessive may also depend on whether deleted exons are alternatively spliced, how many super repeats are deleted, or the involvement of a certain exon or range thereof.

The *NEB* TRI region is included in six of the eigth unique large dominant deletions as well, and it is believed that the *NEB* TRI region, as other segmental duplications and repetitive regions, may mediate the occurrence of these SVs. ^6,52^ The breakpoints of the large deletions coincide mostly with long interspersed nuclear elements (LINE) and Alu elements, but also simple repeats, retrotransposon-originating DNA repeats, and long terminal repeats (LTRs) (Supplementary Table 5, Supplementary Table 6).

Targeted CGH-arrays^16,17^ have been the gold standard methodology for SV detection in *NEB*. To date, it is one of the most robust methods by which the level of mosaicism of large deletions can be determined. Densely targeted custom CGH-arrays allow accurate copy number analysis of the entire gene, including CNVs of the *NEB* TRI. However, accurate copy number estimation of the *NEB* TRI is lost at higher copy numbers due to the method’s binary logarithmic approach. ^17,18^ SVs in *NEB* have been detected by MLPA, panel and/or exome sequencing, and genome short-read sequencing as well. ^21,22^

A targeted ddPCR-based method for *NEB* TRI CNV screening has previously been published, ^18^ allowing for screening in cases where a *NEB* TRI CNV is suspected as a second pathogenic variant, or for segregation analysis in families in which affected individuals have already been shown to be heterozygous for pathogenic CNVs of the *NEB* TRI using other methods (performed for F7).

The downsides of both the published custom CGH-arrays and the ddPCR assays developed for the *NEB* TRI region are that the methods are unable to distinguish between alleles and are limited to analyzing the genomic copy number, i.e. analyzing the expression of gained blocks from cDNA or RNA is not possible using these methods. Secondly, the methods are based on the similarity of the repeated sequences, and they can therefore not distinguish which 8-exon-block is gained or lost, in which orientation gained blocks reside in relation to the gene, or whether duplicated sequences are located within the gene or elsewhere in the genome.

State-of-the-art genomic technologies, such as long-read sequencing (LRS) and optical genome mapping (OGM), show great promise in the detection of SVs in cases where standard-of-care approaches have failed to identify causative variants. ^53,54,55^ Paired with bioinformatic tools especially developed for this purpose, such as Paraphase, LRS may soon also be able to identify *NEB* TRI copy numbers at equal sensitivity, ^56^ with the added advantage of allele phasing.

To date, no inversions, translocations, or larger complex rearrangements have been described in *NEB*. As OGM and LRS become more readily available, these types of SVs may well be identified in previously undiagnosed families.

In summary, we describe a cohort of 23 families with recessive and 12 families from around the world with dominant SVs in *NEB*. This study presents the largest cohort of families (n = 35) to date with pathogenic *NEB* SVs, both recessive and dominant. We establish a clear phenotype-genotype correlation between large dominant deletions of more than 51 exons and distal myopathy.

## Data Availability

All data produced in the present study are available upon reasonable request to the authors.

## Acknowledgements

We thank Marilotta Turunen and Nicolas Dondaine for excellent technical assistance, and MSc Nick Zomer for valuable comments on the manuscript.

## Notes

**Funding statement** This study was supported by the Folkhälsan Research Foundation, the Sigrid Jusélius Foundation, the Medicinska understödsföreningen Liv och Hälsa r.f., Finska Läkaresällskapet, Stiftelsen Perkléns Minne, the Jane and Aatos Erkko Foundation, Muscular Dystrophy UK, and the Waldemar von Frenckells stiftelse. L.S. was supported by a Sigrid Jusélius fellowship (220540). F.H. and K.N. were supported by an MRC strategic award to establish an International Centre for Genomic Medicine in Neuromuscular Diseases (ICGNMD) MR/S005021/1. K.N. received additional Fellowship support from the Guarantors of Brain (UK Charity 1197319). M.S. was supported by the Academy of Finland. The work in C.G.B.’s laboratory is supported by intramural funds from the NIH National Institute of Neurological Disorders and Stroke. Analysis of families F22, F23 and F33 was supported in part by A Foundation Building Strength for Nemaline Myopathy, Boston Children’s Hospital IDDRC Molecular Genetics Core Facility funded by P50HD105351 from the National Institutes of Health of USA, and the Boston Children’s Hospital CRDC Initiative.

**Conflict of Interest statement** The authors declare no conflicts of interest.

### Competing Interest Statement

The authors have declared no competing interest.

### Funding Statement

This study was supported by the Folkhalsan Research Foundation, the Sigrid Juselius Foundation, the Medicinska understodsforeningen Liv och Halsa r.f., Finska Lakaresallskapet, Stiftelsen Perklens Minne, the Jane and Aatos Erkko Foundation, Muscular Dystrophy UK, and the Waldemar von Frenckells stiftelse. L.S. received additional fellowship support from the Sigrid Juselius Foundation (220540). F.H. and K.N. were supported by an MRC strategic award to establish an International Centre for Genomic Medicine in Neuromuscular Diseases (ICGNMD) MR/S005021/1. K.N. received additional Fellowship support from the Guarantors of Brain (UK Charity 1197319). M.S. was supported by the Academy of Finland. The work in C.G.B.'s laboratory is supported by intramural funds from the NIH National Institute of Neurological Disorders and Stroke. Analysis of families F22, F23 and F33 was supported in part by A Foundation Building Strength for Nemaline Myopathy, Boston Children's Hospital IDDRC Molecular Genetics Core Facility funded by P50HD105351 from the National Institutes of Health of USA, and the Boston Children's Hospital CRDC Initiative.

### Author Declarations

The study was performed according to the stipulations of the Declaration of Helsinki of 1975. Written informed consent was obtained from participants and data was de-identified. The study has been approved by the Ethics Committee of the Children's Hospital, University of Helsinki, Finland (approval number 6/E7/05). The approval was renewed by the Ethics Review Board of Helsinki University Hospital in 2021. Where participants were recruited as part of a research project, local ethical approval was obtained: Research Ethics Board of the Hospital for Sick Children (approval number 1000009004), the Human Research Ethics Committee of Stellenbosch University (approval number S22/01/004), the Institutional Review Board of the National Institutes of Health (protocol 12-N-0095), and the Boston Children's Hospital Institutional Review Board (protocol number 03-08-0128R).

