## Supplementary File 1 for "Structural variation in nebulin and its implications on phenotype and inheritance: establishing a dominant distal phenotype caused by large deletions"

### Supplementary file 1: Structural variation in nebulin and its implications on phenotype and inheritance: establishing a dominant distal phenotype caused by large deletions, Sagath et al.

#### Variation in gnomAD

The newest iteration of the SV dataset of the Genome Aggregation Database (gnomAD SVs v.4.1.0, access date April 24 2024) holds records of 68 different intragenic SVs in *NEB*, of which 30 affect one or more exons (Supplementary Table 2).

Of these, 8 are different CNVs of the-*NEB* TRI region, and 22 constitute SVs in the form of duplications and deletions outside the TRI region. In addition to these, the gnomAD lists 38 intronic insertions, deletions, and duplications. Of the SVs affecting exons, 18 are deletions and 12 duplications. Nine of these reside upstream of the *NEB* TRI, and 13 downstream. The average size of the deletions affecting exons is 6,882.8 bp (75 bp to 23.7 kb), with an allele frequency median of 0.00002. The average size of the duplications affecting exons is 5,697.5 bp (75 bp to 21.9 kb), with an allele frequency median of 0.00002. None of the SV records have an allele frequency exceeding 0.01. There is no statistical significance in the difference in size or number of exons included between the deletions and the duplications (Table A, T-test, $p > 0.5$ respectively).

*Table A. Distribution of deletions and duplications affecting NEB exons recorded in gnomAD SVs 4.1.0.*

| SV type | Deletions | Duplications | Difference significance |
|---|---|---|---|
| **Number of unique SVs** | 18 | 12 | |
| **Average number of exons (median, range)** | 4.8 (5, 1-18) | 4.75 (4, 1-17) | p = 0.9598 |
| **Average length (median)** | 6,882.8 (5,389.5) | 5,697.5 (6000) | p = 0.5899 |

GnomAD SVs v4.1.0 holds records of 1035 alleles (34 homozygous) with *NEB* TRI CNVs (Supplementary Table 2). CNVs of the *NEB* TRI region are thus the most common variant in *NEB*, with a combined allele frequency of 0.009. The prevalence of CNVs in the *NEB* TRI region renders it a true mutational hotspot. These *NEB* TRI CNVs are thought to occur by non-allelic homologous recombination (NAHR) mediated by the long interspersed nuclear elements (LINE) riddled in the introns of the repeated region.[6,57,58]

#### Statistical tests

All statistical tests were performed in RStudio 2022.02.1 build 461.

##### *Correlation between gene length and number of recorded SVs*

The genomic gene lengths and number of structural variants were extracted recorded in gnomAD SVs 4.1.0 for 12 known and well-established NM-causing genes (Supplementary table 3) and *MYO18B*. The gene length was extracted based on the first and last basepair of the respective representative transcripts from RefSeq & GENCODE (Matched Annotation from NCBI and EMBL-EBI, MANE Select Plus Clinical) were used. The gene length was plotted against the number of recorded SVs using ggplot2 to verify normal distribution (Figure A). The Pearson correlation and Spearman correlation coefficients and corresponding p-values were calculated.

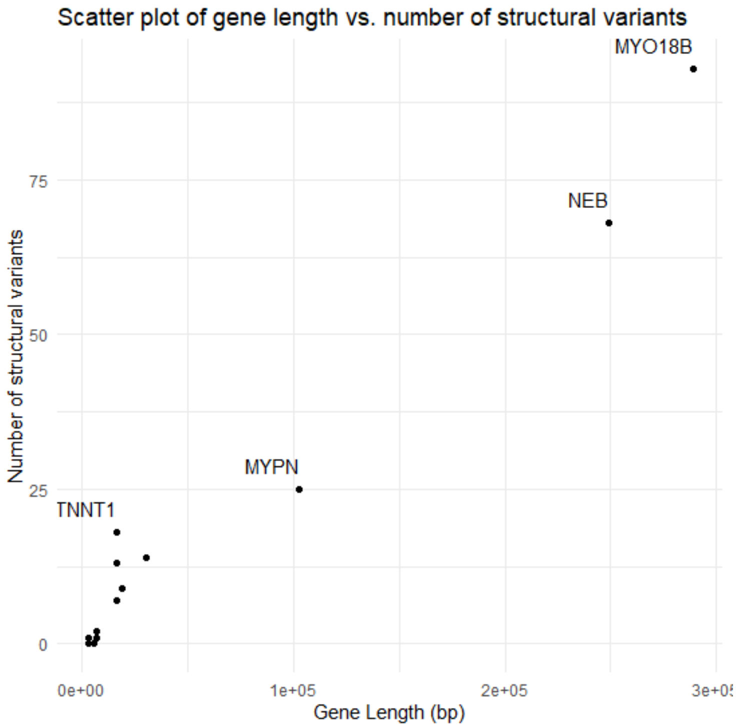


*Figure A. Scatter plot showing the gene length in base pairs plotted against the number of structural variants recorded in the respective genes in gnomAD SVs 4.1.0.*

The Pearson correlation coefficient was 0.9821 ($p = 2.534 * 10^{-9}$), and the Spearman correlation coefficient was 0.9254 ($p = 5.722 * 10^{-6}$).

In gnomAD SVs v.4.1.0 the number of SVs in *NEB* is thus higher than in any other well-established NM-causing gene (Supplementary Table 3). Based on the correlation analysis, this can be attributed to the size of the gene, as the size of the NM-causing genes directly correlates with the number of recorded intragenic SVs in them.

##### *Correlation between different types of small variants and phenotype*

Families with recessive variants and enough phenotypic data were classified into three groups: mild, typical, and severe, according to established classification criteria (Wallgren-Pettersson et al. 2000). Small variants were divided into splicing, missense and truncating variants, of which the latter includes both nonsense and frameshift variants. Families with homozygous *NEB* SVs (F20) and TRI CNVs (F12), and families in which severity classification was not possible (F2, F5, F13, F17, F19) were excluded from the statistical test. Patients were divided into two groups: those with *NEB* TRI CNVs in trans with a small variant (CNV-SNV) and those with other SVs in trans with a small variant (SV-SNV). The input data is shown in Table B.

*Table B. Input data for the correlation analysis between different types of small variants and phenotype.*

| Family | Category | SVExonCount | TRICNPath | SNVExon | Class | Severity |
|---|---|---|---|---|---|---|
| F1 | CNV-SNV | NA | 7 | 3 | Truncating | 3 |
| F3 | CNV-SNV | NA | 7 | 28 | Truncating | 3 |
| F4 | CNV-SNV | NA | 11 | 59i | Splice | 2 |
| F6 | CNV-SNV | NA | 6 | 78 | Inframedel | 2 |
| F7 | CNV-SNV | NA | 7 | 81 | Truncating | 2 |
| F8 | CNV-SNV | NA | 7 | 126 | Truncating | 2 |
| F9 | CNV-SNV | NA | NA | 157 | Truncating | 2 |
| F10 | CNV-SNV | NA | 6 | 134 | Truncating | 1 |
| F11 | CNV-SNV | NA | 6 | 180 | Truncating | 2 |
| F14 | SV-SNV | 51 | NA | 78 | Truncating | 1 |
| F15 | SV-SNV | 24 | NA | 129 | Missense | 3 |
| F16 | SV-SNV | 3 | NA | 155 | Truncating | 2 |
| F18 | SV-SNV | 2 | NA | 36i | Splice | 2 |
| F21 | SV-SNV | 1 | NA | 173 | Truncating | 2 |
| F22 | SV-SNV | 2 | NA | 173 | Truncating | 2 |
| F23 | SV-SNV | 18 | NA | 169 | Truncating | 1 |

The correlation between small variant type and severity of disease was assessed in both patient groups (CNV-SNV and SV-SNV) together, as well as separately.

For the combined analysis, the data was formatted into contingency tables in R by variant class (Class) and severity of the phenotype (Severity). A $X^2$ was performed to assess the expected frequencies for both groups independently and for the combined data. This resulted in expected frequencies under 5, a *post hoc* Fisher's Exact Test for Count Data was used. For the patient group independent tests, only the respective data was used.

Fisher's Exact Test for Count Data for both recessive patient groups yielded a p-value of 0.7405. Fisher's Exact Test for Count Data for the CNV-SNV group yielded a p-value of 1. Fisher's Exact Test for Count Data for the SV-SNV group yielded a p-value of 0.2143.

In either of the groups alone, nor combined, could a statistical significance between the type of small variant and the severity of the disease be established.

##### *Correlation between pathogenic NEB TRI CNV allele and phenotype*

In families with pathogenic gains of the *NEB* TRI region (F1, F3, F4, F6, F7, F8, F10, F11), the correlation between the copy number of the *NEB* TRI of the pathogenic allele and the severity of phenotype was tested using Kendall's Tau. The input data is shown in Table B above, but F9 was excluded as the copy number of the pathogenic allele was not known.

Kendall's rank correlation tau yielded a z-value of 1.133 and a p-value of 0.2572.

No statistical significance between the copy number of the *NEB* TRI and the severity of the disease could be established.

##### 2.5.4. Difference between deletion length between families with NM and distal phenotypes

The length of all unique deletions, both in recessive NM families (n = 7) and dominant distal families (n = 8), were extracted both in number of affected exons and number of base pairs deleted. For deletions in which the exact basepair level breakpoints were unknown, the average length based on the CGH-array variant call was calculated. The input data is shown in Table C.

*Table C. Input data for the correlation analysis between the exon number and variant length for deletions in recessively inherited NM and distal myopathy families.*

| Variant | ExonCount | LengthBp | Group |
|---|---|---|---|
| 1 | 24 | 58523 | NM |
| 2 | 3 | 2250 | NM |
| 3 | 2 | 1083 | NM |
| 4 | 3 | 4193 | NM |
| 5 | 1 | 913 | NM |
| 6 | 2 | 2506 | NM |
| 7 | 18 | 24763.5 | NM |
| 8 | 97 | 139610.5 | Distal |
| 9 | 76 | 102258 | Distal |
| 10 | 64 | 87723 | Distal |
| 11 | 67 | 93022 | Distal |
| 12 | 75 | 103788 | Distal |
| 13 | 94 | 122144 | Distal |
| 14 | 68 | 92335.5 | Distal |
| 15 | 51 | 72281.5 | Distal |

First, data normality was assessed using the Shapiro-Wilk test. The results of these are displayed in Table D.

*Table D. Shapiro-Wilk test results for the two groups versus number of exons deleted and deletion length in bp.*

| Group | TestVariable | W | p |
|---|---|---|---|
| NM | ExonCount | 0.71008 | 0.004771 |
| NM | LengthBp | 0.67092 | 0.00177 |
| Distal | ExonCount | 0.93687 | 0.5806 |
| Distal | LengthBp | 0.951 | 0.7213 |

The equality of variances was assessed *post hoc* using Levene's test for Homogeneity of Variance by exon number and group, and length in bp and group. The results of these tests are displayed in Table E.

*Table E. Levene's test for Homogeneity of Variance between the two groups versus number of exons deleted and deletion length in bp.*

| Group | TestVariable | Df (residual) | F value | Pr(>F) |
|---|---|---|---|---|
| NM/Distal | ExonCount | 1 (13) | 1.4995 | 0.2425 |
| NM/Distal | LengthBp | 1 (13) | 0.1418 | 0.7125 |

Based on the Shapiro-Wilk test, deletion lengths measured both in basepairs and number of exons is not normally distributed in the NM group (significant), but is in the Distal group (not significant). Levene's test, however, indicates that there is no significant difference between the variances in the two groups for either parameter (exon count or length in bp). Hence, we used the Wilcoxon Rank-Sum test (a.k.a. Mann-Whitney U test, non-parametric), which does not assume normality of the data. The result of these are presented in Table D.

*Table D. Wilcoxon Rank-Sum test results for comparison of the distribution of number of exons deleted and deletion length in bp in the two groups.*

| Group | TestVariable | W | p-value |
|---|---|---|---|
| NM/Distal | ExonCount | 56 | 0.001432 |
| NM/Distal | LengthBp | 56 | 0.0003108 |

Based on the Wilcoxon Rank-Sum test, the difference in deletion length, measured either in number of exons deleted or length in basepairs, is significant ($p < 0.05$).
